# A chronopharmacology-friendly multi-target therapeutics based on AI: the example of therapeutic hypothermia

**DOI:** 10.1101/2022.03.07.22271997

**Authors:** Fei Liu, Xiangkang Jiang, Mao Zhang

**Affiliations:** Department of Emergency Medicine, Second Affiliated Hospital of Zhejiang University, Hangzhou 310009, Zhejiang Province, China; Institute of Emergency Medicine, Zhejiang University, Hangzhou 310009, Zhejiang Province, China; Key Laboratory of The Diagnosis and Treatment of Severe Trauma and Burn of Zhejiang Province, Zhejiang University, Hangzhou 310009, Zhejiang Province, China

**Keywords:** multi-target therapeutics, chronopharmacology, precision medicine, Therapeutic hypothermia, virtual screening

## Abstract

Nowadays, the complexity of disease mechanisms and inadequacy of single-target therapies in restoring the biological system has inevitably instigated the strategy of multi-target therapeutics with the application of hybrid and chimeric drugs. However, the related method is still unable to solve the conflicts between targets or between drugs. With the release of high-precision protein structure prediction artificial intelligence (AI), large-scale high-precision protein structure prediction and docking become possible. In this article, we propose a multi-target drug discovery method. Then we take an example of therapeutic hypothermia (TH). We performed protein structure prediction for all targets of each group by AlphaFold2 and RoseTTAFold. QuickVina 2 is then used for molecular docking of the proteins and drugs. After docking, we use PageRank to get the rank of drugs and drug combinations of each group. Given the differences in the scoring of different proteins, the method can effectively avoid inhibiting beneficial proteins. So it’s friendly to chronopharmacology. This method also have potential in precision medicine for its high compatibility with bioinformatics.

## Introduction

At present, the common strategy of drug development is derived from the paradigm “One Drug, One Disease”. Therefore, highly selective and potent molecules are being developed for a distinct target.[1] However, in the application of this strategy, the hybrid drugs or chimeric drugs could not solve the problem of interaction between drugs or targets.[2] Besides, this approach also has disadvantages when dealing with time-specific proteins. These defects limits the application of this strategy.

Take therapeutic hypothermia as an example. Therapeutic hypothermia (TH) can limit the degree of some kinds of injuries in randomized trials[3] and animal experiments[4], and is even the only effective method for some diseases especially hypoxic-ischemic encephalopathy (HIE). HIE often causes severe neurological sequelae, which is the main reason for the poor prognosis of patients with stroke, shock, carbon monoxide poisoning, cerebral hemorrhage, and cardiac arrest.[5–7]

In the research based on TH, cold shock proteins especially cold-induced RNA binding protein (CIRP) show high expression [8] and rapid response [9]. CIRP has been shown to promote the translation of genes involved in DNA repair [10,11], telomerase maintenance[12], and genes associated with the translational machinery[13].

However, if CIRP leaks to the intercellular substance with cell swelling and rupture, it will become a harmful protein. Extracellular CIRP (eCIRP) showed a strong pro-inflammatory effect, leading to a heavier hypoxic injury.[14,15] So, agonists applied with CIRP can effectively promote cell protection before CIRP leaks out of the cell. After the leakage of CIRP out of the cell, the application of the CIRP antagonist can effectively promote cell protection.

With the development of high-precise protein prediction technologies, especially AlphaFold2[16] and RoseTTAFold[17], we can obtain the structure of proteins quickly and accurately, and widespread docking becomes possible. In this article, we provide a multi-targeted drug discovery method. This method attempts to include all proteins as targets for analysis. For all proteins, antagonize the negative proteins as much as possible without affecting the positive ones.

## Method

### Experiment design

As shown in Figure 1, the representative experiment is divided into 5 processes: 1. Protein targets were calculated by bioinformatics analysis; 2. Protein and drugs 3D structure acquisition and prediction; 3. Proteins’ active sites prediction; 4. Drug/molecular group evaluation with target proteins; 5. PageRank of docking results, protein logFC, and mRNA expression. The experiment of animals or cells is referred to by authors, but not forced. The biggest difference from previous studies is PageRank.

**Figure 1.**
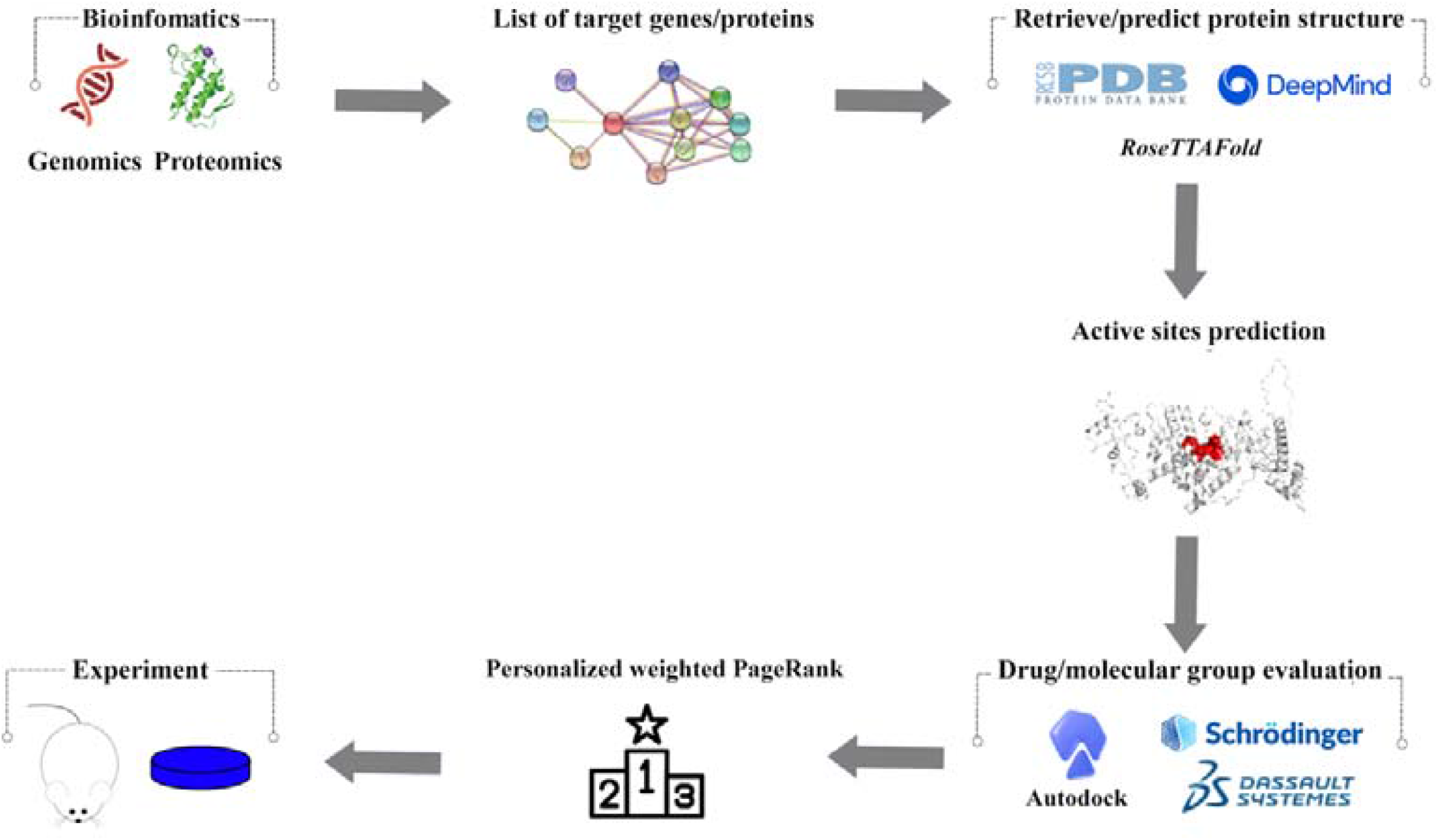
Representative workflow for bioinfo-pharmacology drug design.

### The data source of bioinformatics analysis

We retrieved the original data of mRNA expression under hypothermia treatment from the website of The National Center for Biotechnology Information (NCBI) (GSE54229). The research was reported by Sten et.al.[9] In their research, mouse embryonic fibroblasts were exposed to mild hypothermia (32°C) or normothermia (37°C) to gain the transcription response induced by hypothermia. After 0.5, 1, 2, 4, 8, and 18 hours of hypothermia, cells were collected for bioinfomatics analysis.

### Expression Profile Analysis

The log2 fold-change (log2FC) and p-value were calculated for the normothermia group. Top 3 log2FC mRNA with q-value < 0.05 were selected from each group to enter the next step. If there exists mRNA with failed protein structure prediction, the mRNA would be skipped.

R 3.6.1 was used to detect di□erential expressed compared to matched normothermia samples. The clustering of genes was calculated by the “dist” and “hclust” function of R. The visualization of gene expression and clustering is performed by the “dendextend” package.

### 3D Data of proteins and small molecular drugs

All proteins were first searched on PubMed to see if there was protein clipping like cleaved caspase-3[18].

Then the 3D structures of proteins were firstly searched from Protein Data Bank (PDB), which is used for biological-related ligand-protein interaction. In this article, no protein structure is listed on the PDB website. All the protein structures were predicted by AlphaFold2 and RoseTTAFold.

AlphaFold2 is developed by Google and is the champion of the 14^th^ Critical Assessment of Structure Prediction (CASP14). In August 2021, AlphaFold submitted a structure prediction database for all proteins. RoseTTAFold is based on the Rosetta software which is designed for macromolecular modeling, docking, and design[19] RoseTTAFold also has good application[20] in the research of protein structure prediction. Finally, protein structures with fewer irregular regions will be selected for the next step.

The 3D structures of 8,697 drugs (DrugBank, 5.1.8) were downloaded from DrugBank Online (https://go.drugbank.com/). Approved, experimental, nutraceutical, and investigational drugs by Food and Drug Administration (FDA) are included. We split each drug molecule into a PDBQT-format file and minimized the energy separately for docking with proteins.

### Visualize evolutionary conservation and active site prediction

Visualize evolutionary conservation was performed by the ConSurf server[21]. In a typical ConSurf application, through BLASTed[22] against the UNIREF-90 database[23] and aligning using MAFFT[24], the evolutionarily conserved positions are analyzed by the Rate4Site algorithm.

Then, the Consensus approach-D (COACH-D) [25] was used to predict the active site of target proteins. The COACH-D uses five different methods to predict the binding sites of protein ligands. Four of these methods are COFAC-TOR[26], FINDSITE[27], TM-SITE[28], and S-SITE[28]. These methods predict binding sites by matching the query structure and sequence with the ligand-binding template in BioLiP[29], which is a semi-manual functional database[30] based on the PDB.

### Virtual screening of potential compounds

To evaluate the hit compounds obtained from DrugBank and calculate their interaction and binding posture in the active site of target proteins, the molecular docking method was carried out through QuickVina 2[31]. QuickVina 2 uses the calculation of shape and electrostatic potential similarity of binding pockets to select molecules, which may exhibit binding patterns like those of binding pockets.

3D files of target proteins were dehydrated and hydrogenated. Then proteins were saved as PDBQT files using AutoDock. AutoDock assisted in assigning Gasteiger charges and adding polar hydrogen atoms to both the proteins and the compounds.

### Molecular dynamics simulation

The molecular dynamics (MD) simulation was performed by Gromacs[32]. Firstly, a protein-drug complex was prepared, including adding hydrogenation and balancing charge. Then, we add a solvent so that the target protein and small molecules are coated. The forcefield was Chemistry at HARvard Macromolecular Mechanics 36 (CHARMm 36). The simulation time is set as 50ns for the speed of calculation. The simulation temperature is 309.15K (36□) and the pressure is 1 atm. Root mean square deviation (RMSD) and root mean square fluctuation (RMSF) were calculated based on the first frame.

### Personalization-weight-PageRank

We use personalization-weight-PageRank to rank cross-level data. PageRank is a comprehensive rank algorithm designed by Google and named after Larry Page.[33] It is one of the most famous ranking algorithms of network nodes based on the Markov process. PageRank has been applied in medical domains with success.[34,35]

Personalization and weight represent 3 different levels of score data. The weight of PageRank allows all nodes to be initially assigned different weights/probabilities.[36] In this article, the weights of rank were set to docking values of proteins and drugs. The higher the docking value, the higher the connection rate of the complex.

Personalization of PageRank reinforces the connection intensity between the nodes, which makes the result more personalized and realistic[37]. In this article, personalization is influenced by protein functions. If the protein performs a negative influence such as promoting apoptosis, the personalization will be calculated by 2^(fold change) to ensure they are more than 1. Meanwhile, if the protein plays a positive role in the group, the personalization will be set as 1/(fold change + 1) to less than 1. The personalization values of all the drugs are set to 0 to prevent iterations of the drugs themselves from going wrong.

The calculation process is like putting all proteins and all drugs in the solution, then simulating the connections between all proteins and drugs by calculation. The damping factor is set to 0.85 to simulate the metabolism of proteins and drugs.

The whole calculation is based on Python 3.8.10. The relating python libraries include NetworkX, Pandas, and NumPy. We use Pandas and NumPy to import all the docking data into a matrix for PageRank calculating. The protein expression value is then imported by the PageRank personalization parameter of NetworkX. Lastly, we can get a comprehensive ranking of drugs.

### Prediction and Rank of combined pharmacotherapy

In addition to the comprehensive ranking of drugs, we also try to generate the rank of drug combinations. Similarly, the calculation places all drugs of combination and target proteins in a solution to bind free.

First, all drugs will be grouped according to the docking results of drugs in each combination. In this article, to reduce the amount of calculation, we selected the TOP20 drugs of each protein to include in the drug combination pool. Then, all the combinations were performed personalization-weight-PageRank against all protein targets. The sum of each score of all drugs in the combination is the final score of the combination. Lastly, we get the rank of combinations.

To make the distribution of combinations clear, we propose drug-protein-expression fit score (DPEFS) to show the data distribution pattern. The calculation is as follows: The PageRank values of all proteins were summed by multiplying logFC, then divided by the total PageRank values of drugs, and finally divided by the PageRank values of specific proteins for standardized calculation. It is used for standardized calculation for comparing different combinations.

DPEFS evaluates the combination by referring to the protein expression trend. The higher the DPEFS, the better the fitness. In actual drug design, DPEFS is relatively high and PageRank score is relatively low, indicating that drugs of combination are relatively moderate, which suggests a negative outcome. All code can be found on GitHub (https://github.com/FeiLiuEM/PageRank-weight-drug).

## Result

### Expression analysis and clustering of hypothermia

Figure 2 shows the expressions of different mRNA of different groups after hypothermia. From the inside to the outside, the rings were divided into hypothermia 0.5h group, hypothermia 1H group, hypothermia 2H group, hypothermia 4H group, hypothermia 8h group, and hypothermia 18h group.

**Figure 2.**
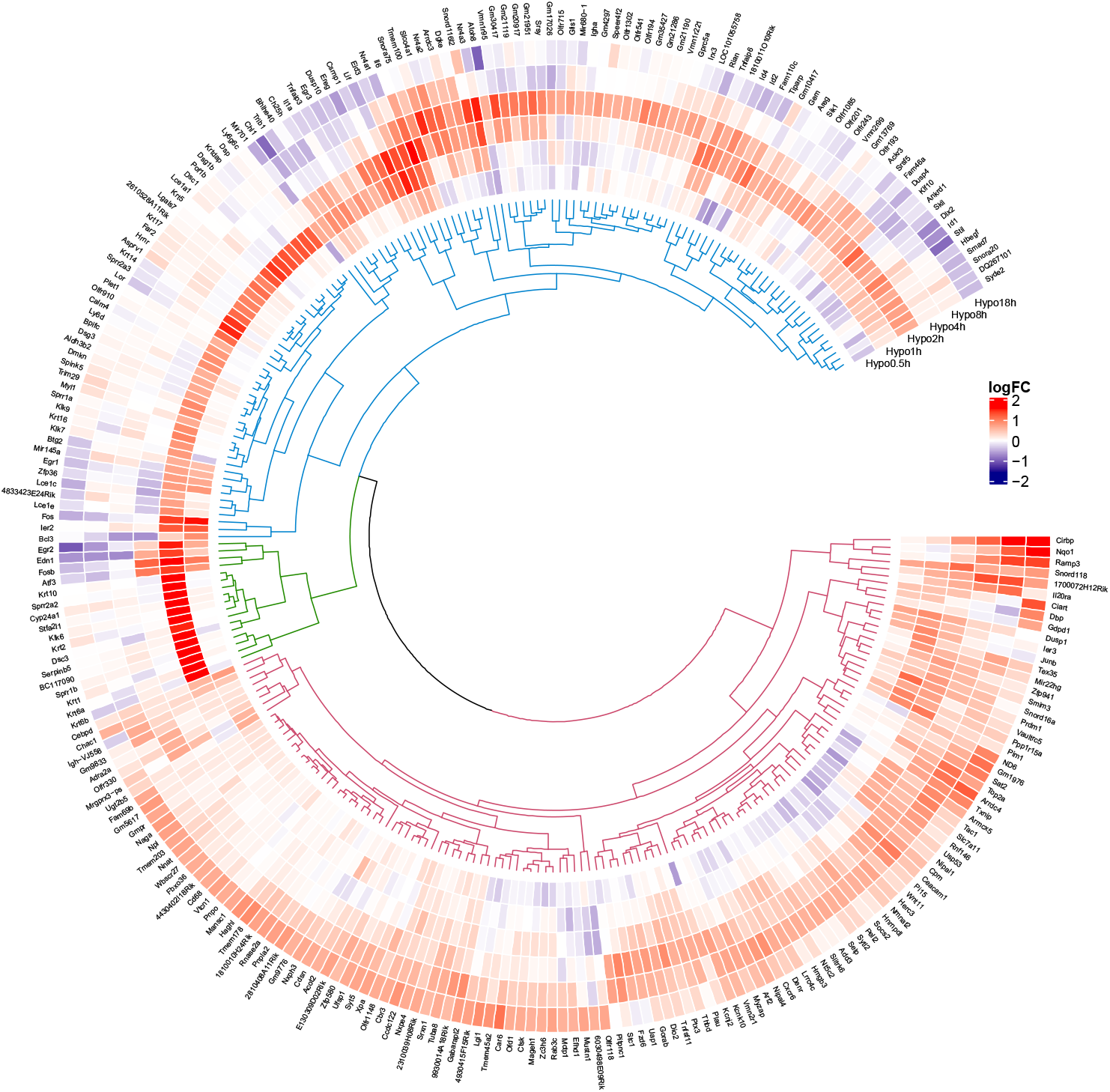
Circular visualization of expression patterns and clustering of hypothermia treatment. Red indicates gene upregulation and blue indicates downregulation.

As shown in Table 1, in each group, we selected the top 3 expression protein targets. In the Hypothermia 0.5h group, the target proteins are circadian-associated transcriptional repressor (CIART), Glutathione-specific gamma-glutamylcyclotransferase 1 (CHAC1), and Uridine diphosphate glucose pyrophosphatase nudix hydrolase 22 (NUDT22). The target proteins of the Hypothermia 1h group are CHAC1, corneodesmosin (CDSN), and Nuclear receptor subfamily 1 group D member 1 (NR1D1). The target proteins of the Hypothermia 2h group are cold-induced RNA-binding protein (CIRP), armadillo repeat-containing X-linked protein 5 (ARMCX5), and coiled-coil domain-containing protein 122 (CCDC122). The target proteins of the Hypothermia 4h group are CIRP, receptor activity-modifying protein 3 (RAMP3), and carcinoembryonic antigen-related cell adhesion molecule 1 (CEACAM1). The target proteins of the Hypothermia 4h group are the same: CIRP, RAMP3, and NAD(P)H dehydrogenase [quinone] 1 (NQO1).

**Table 1.**
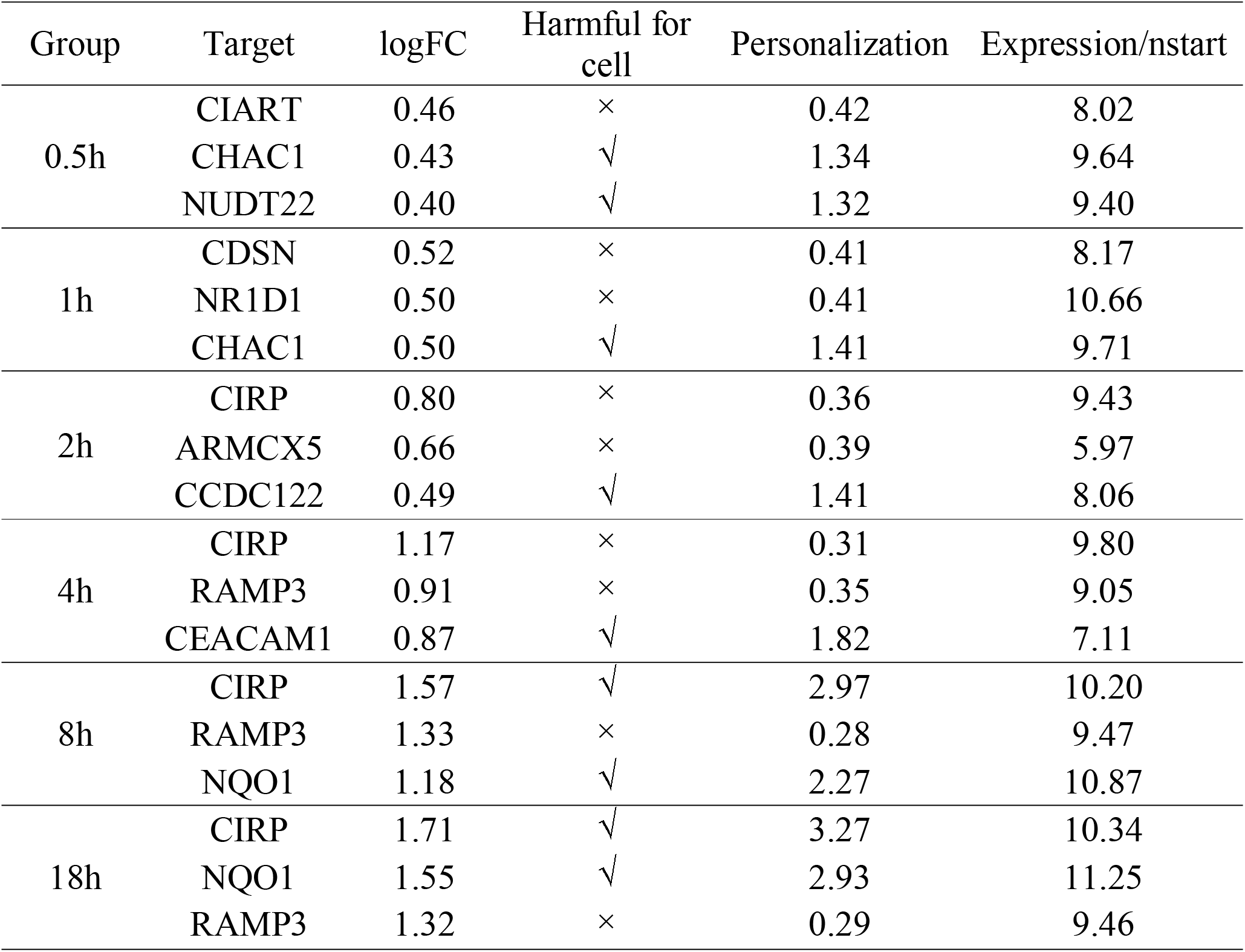
The target proteins of different groups.

Within the targets, CHAC1 could enhance apoptosis[38]. NUDT22 is an Mg^2+^-dependent UDP-glucose and UDP-galactose hydrolase[39], while high glucose shows a negative effect in HIE like stroke[40]. CCDC122 potentially pro-inflammatory[41]. CIRP can effectively reduce cell death in the early stage of hypothermia therapy. However, it has a strong pro-inflammatory effect outside the cell, leading to cell killing. There is no definitive research on the timing of this shift. Referring to the previous article[42], we conservatively believed that CIRP could be identified as a negative protein from the 8H group. CEACAM1[43] and NQO1[44] promote apoptosis. All the other targets are shown protective effects or don’t have enough data. The personalization values were calculated in Table 1. All the structures of target proteins in Figure 3 were obtained by the rules in the section of Materials and Methods.

**Figure 3.**
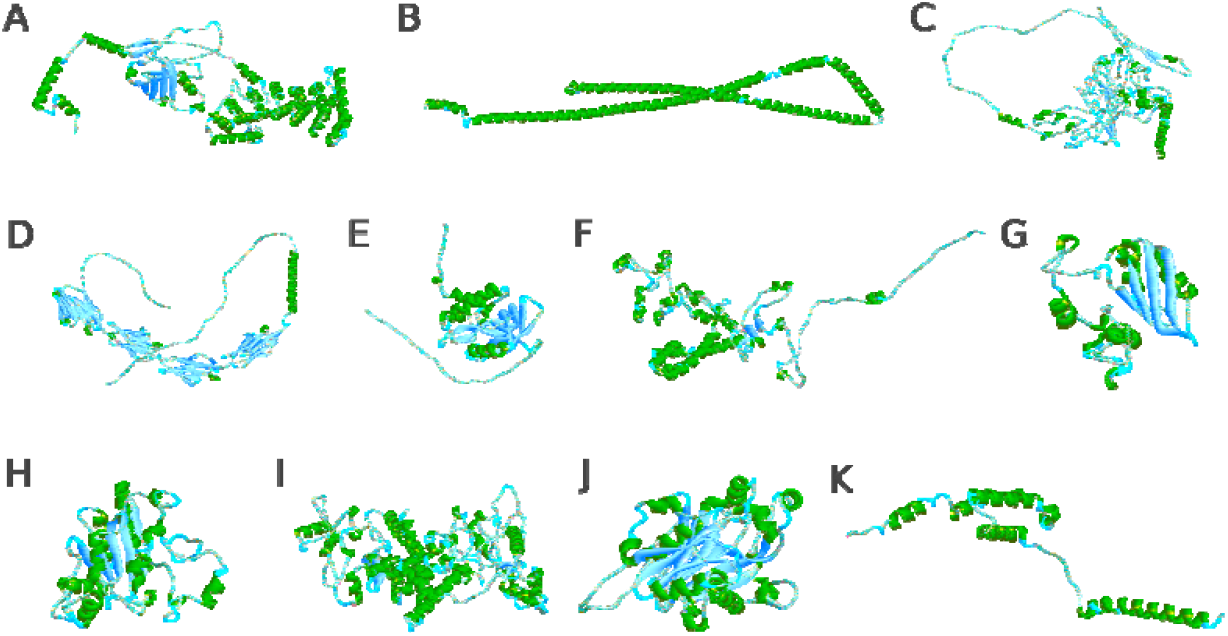
The 3D structures of target proteins. A. ARMCX5; B. CCDC; C. CDSN; D. CEACAM1; E. CHAC1; F. CIART; G. CIRP; H. NQO1; I. NR1D1; J. NUDT22; K. RAMP3.

### Visualize evolutionary conservation and Structure-Function Relationship-Based Binding Site Prediction

The conservation analysis of all the target proteins was listed in Figure 4A-K. The redder the amino acid, the higher possibility of the amino acid sequence with function. Then we identified its structure-function relationship by the COACH-D server. The results showed a familiar result of conservation analysis listed in Figure 4L-V. As shown in Table 2, the range around 3-5 Å of the active site was used for the setting of the receptor pocket of the target proteins that were used for virtual screening.

**Figure 4.**
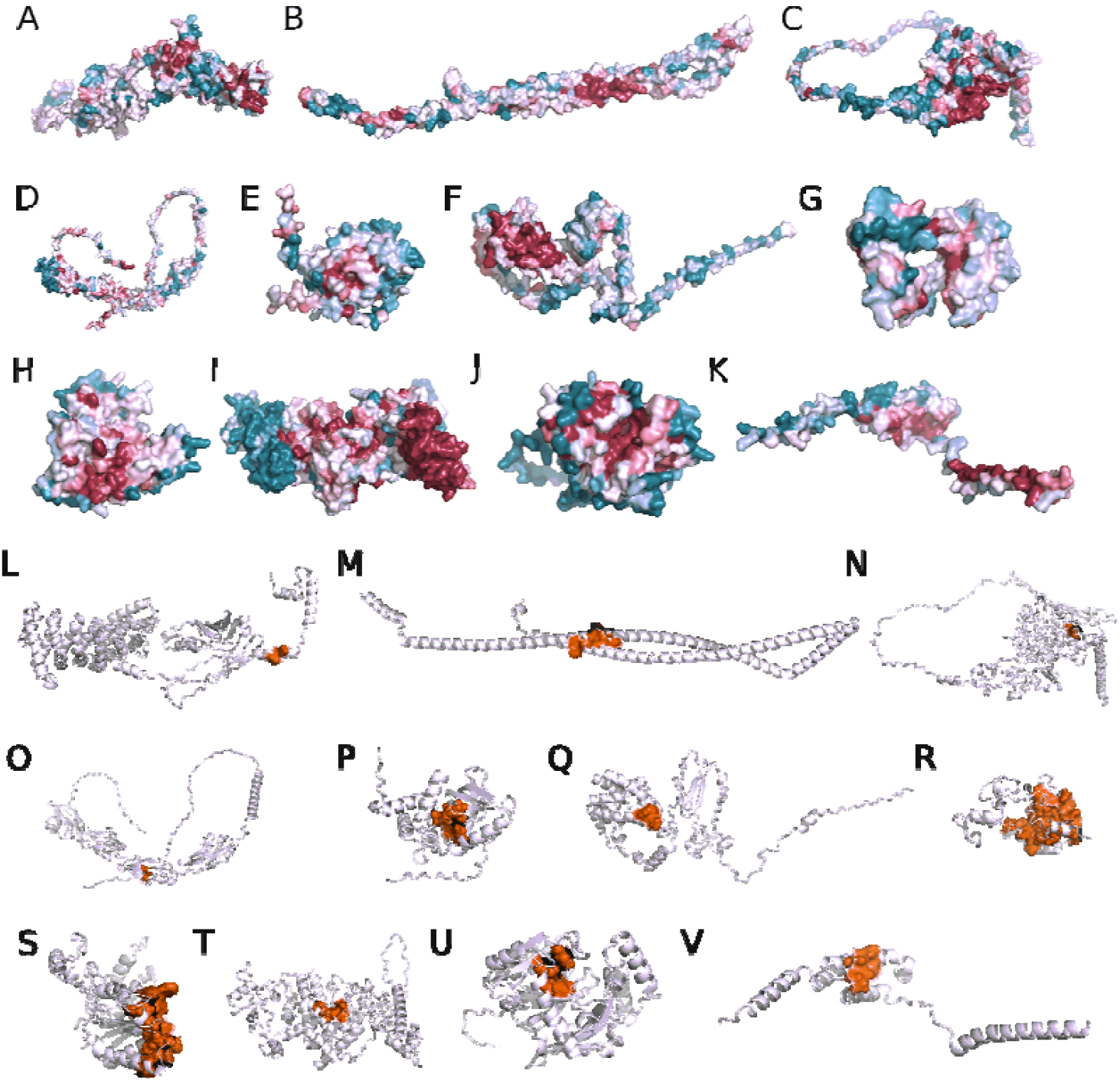
The ConSurf analysis and predicted active sites of target proteins. The upper 11 pictures are ConSurf analysis results. The last 11 pictures are predicted active sites. A&L. ARMCX5; B&M. CCDC; C&N. CDSN; D&O. CEACAM1; E&P. CHAC1; F&Q. CIART; G&R. CIRP; H&S. NQO1; I&T. NR1D1; J&U. NUDT22; K&V. RAMP3. The redder the amino acid, the more conservative it is. The greener the color, the less conservative it is.

**Table 2.** The docking parameters of target proteins.

| Protein name | protein source | X | Y | Z | LEN-X | LEN-Y | LEN-Z |
| --- | --- | --- | --- | --- | --- | --- | --- |
| ARMCX5 | RoseTTAFold | -7.06 | 31.20 | -43.05 | 29.25 | 28.50 | 24.75 |
| CDSN | RoseTTAFold | 51.80 | 53.51 | 31.17 | 39.75 | 20.25 | 21.00 |
| CEACAM1 | AlphaFold2 | -41.13 | 3.42 | 5.67 | 39.75 | 22.50 | 22.50 |
| CHAC1 | AlphaFold2 | -3.04 | -0.53 | 0.22 | 23.25 | 25.50 | 35.25 |
| CIART | RoseTTAFold | 36.79 | -16.59 | -36.23 | 28.50 | 38.25 | 22.50 |
| CIRP | RoseTTAFold | 14.48 | 2.86 | -9.56 | 17.25 | 15.00 | 19.50 |
| NQO1 | AlphaFold2 | -1.86 | -16.32 | -9.20 | 29.25 | 29.25 | 28.50 |
| NR1D1 | RoseTTAFold | 0.34 | 35.70 | 10.28 | 39.75 | 25.50 | 24.75 |
| NUDT22 | AlphaFold2 | 10.08 | -12.04 | 14.09 | 17.25 | 38.25 | 22.50 |
| RAMP3 | AlphaFold2 | 0.00 | -10.93 | -5.29 | 24.00 | 24.00 | 21.00 |
| CCDC122 | RoseTTAFold | 119.27 | 23.34 | 8.09 | 36.00 | 47.25 | 47.25 |

### Virtual Screening of target proteins’ Antagonists

We utilized the virtual screening technique to identify potential antagonists exhibiting an adequate binding a□nity. We started with a chemical database consisting of 8,697 drug molecules and isolated a set of compounds satisfying the threshold of a high docking score. The results of the best match complexes are shown in Figure 5 and all the results are listed in the Additional file Table 1.

**Figure 5.**
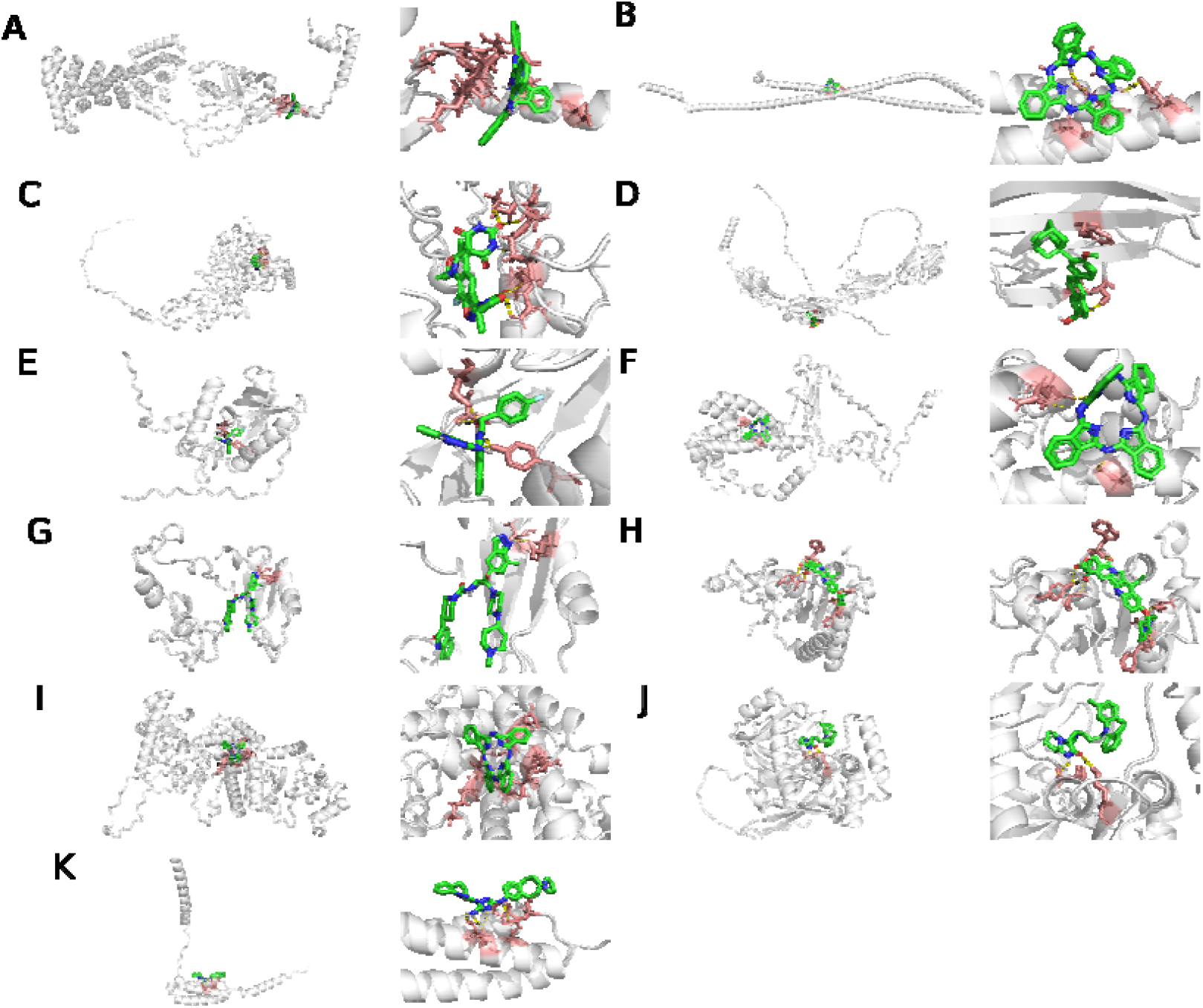
The best docking molecular for each protein. A. ARMCX5; B. CCDC; C. CDSN; D. CEACAM1; E. CHAC1; F. CIART; G. CIRP; H. NQO1; I. NR1D1; J. NUDT22; K. RAMP3.

### MD Simulations and Binding Free Energy Analysis

We performed the MD simulation of 11 complexes to measure the stability of the protein-ligand complex. RMSD (root-mean-square deviation) profiles of the protein are shown in Figure 6A, which indicates that all systems were relatively stable during the entire simulation run. Moreover, RMSF profiles of protein are measured to evaluate the moving of each amino acid. All proteins are available for further analysis (Figure 6B).

**Figure 6.**
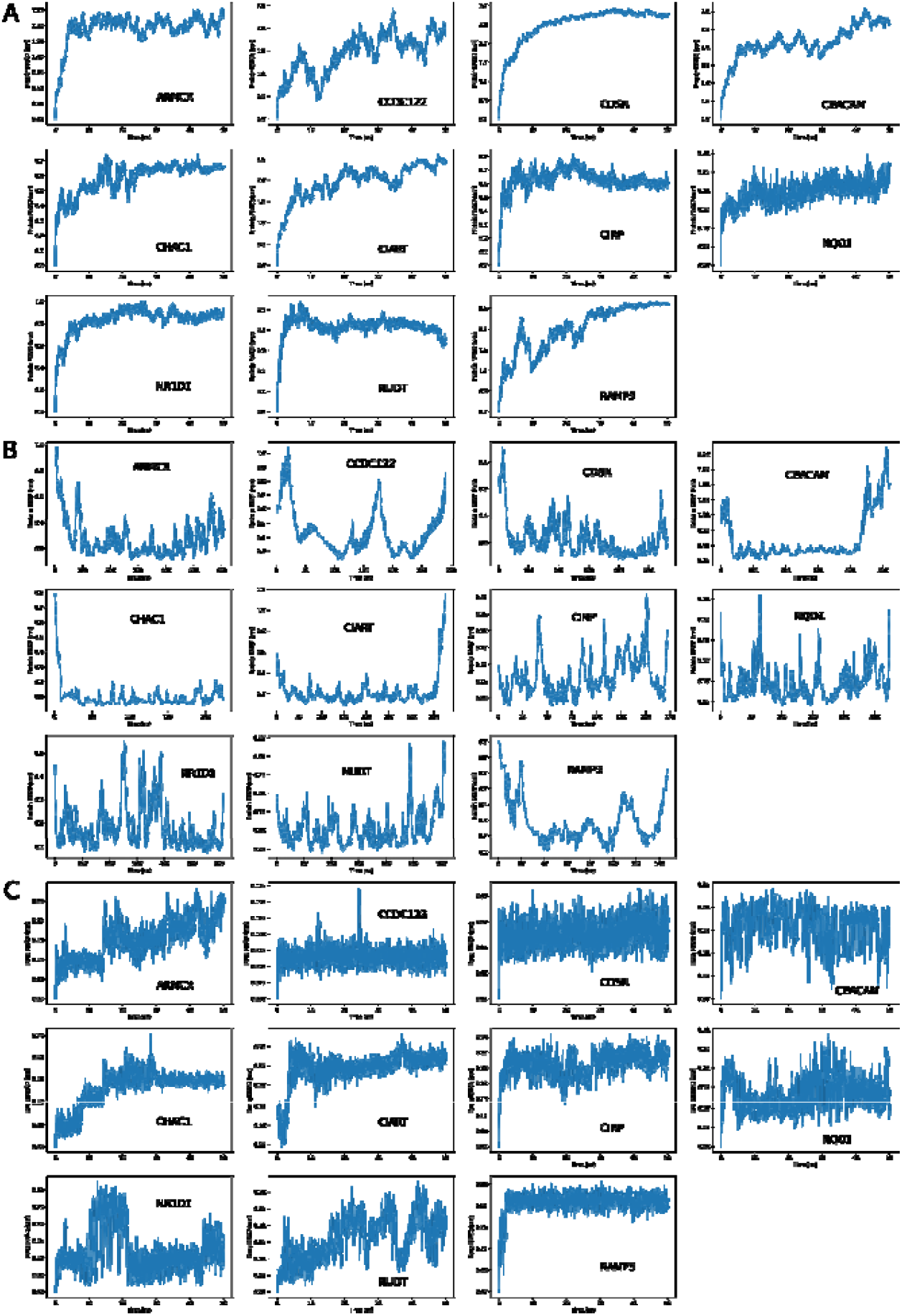
The RMSD and RMSF of MD simulation. A. The RMSD of proteins. B. The RMSF of proteins. C. The RMSD of each molecular of proteins.

The RMSD of drug atoms was also conducted to predict the stability of the atoms in docked complexes (Figure 6C). Most compounds exhibited a consistently low RMSD, suggesting that these compounds formed stable complexes.

### Drug rank of TH in different groups

We rank all drugs by PageRank. First, we PageRank all the drugs and get the results in table3. The higher the rank, the protective effect in TH. The lower the rank is, the more damaging it is in TH.

**Table 3.**
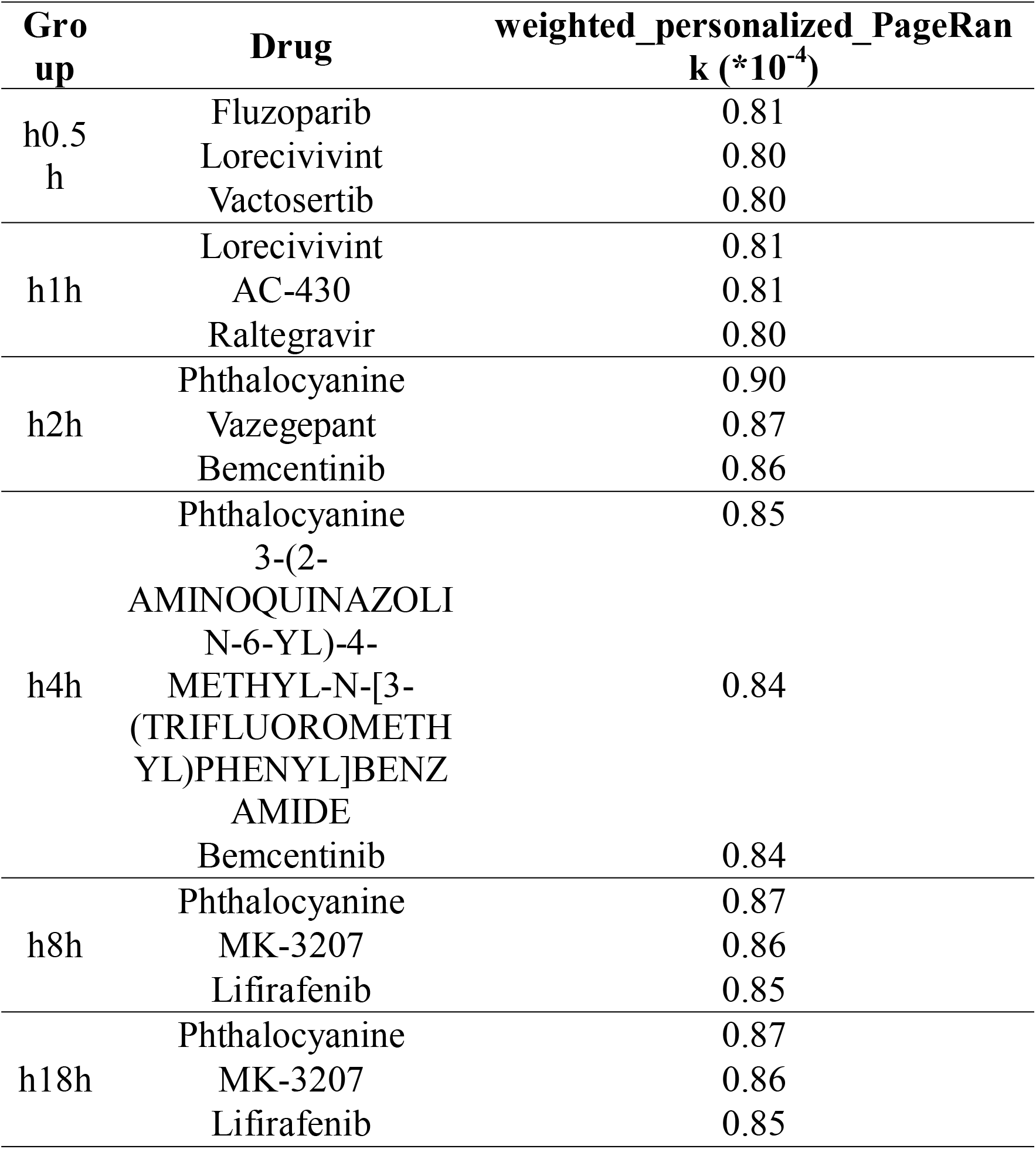
The comprehensive rank of drugs at different groups.

2-drug-combinations are ranked in Table 4 and 3-drug-combinations in the additional file Table 2. For comprehensive rank, the results of PageRank were listed. For drug-combination ranks, the percentages of each drug’s value in the combination were calculated. And DPEFS was calculated for analyzing the distribution differences of drug combinations.

**Table 4.**
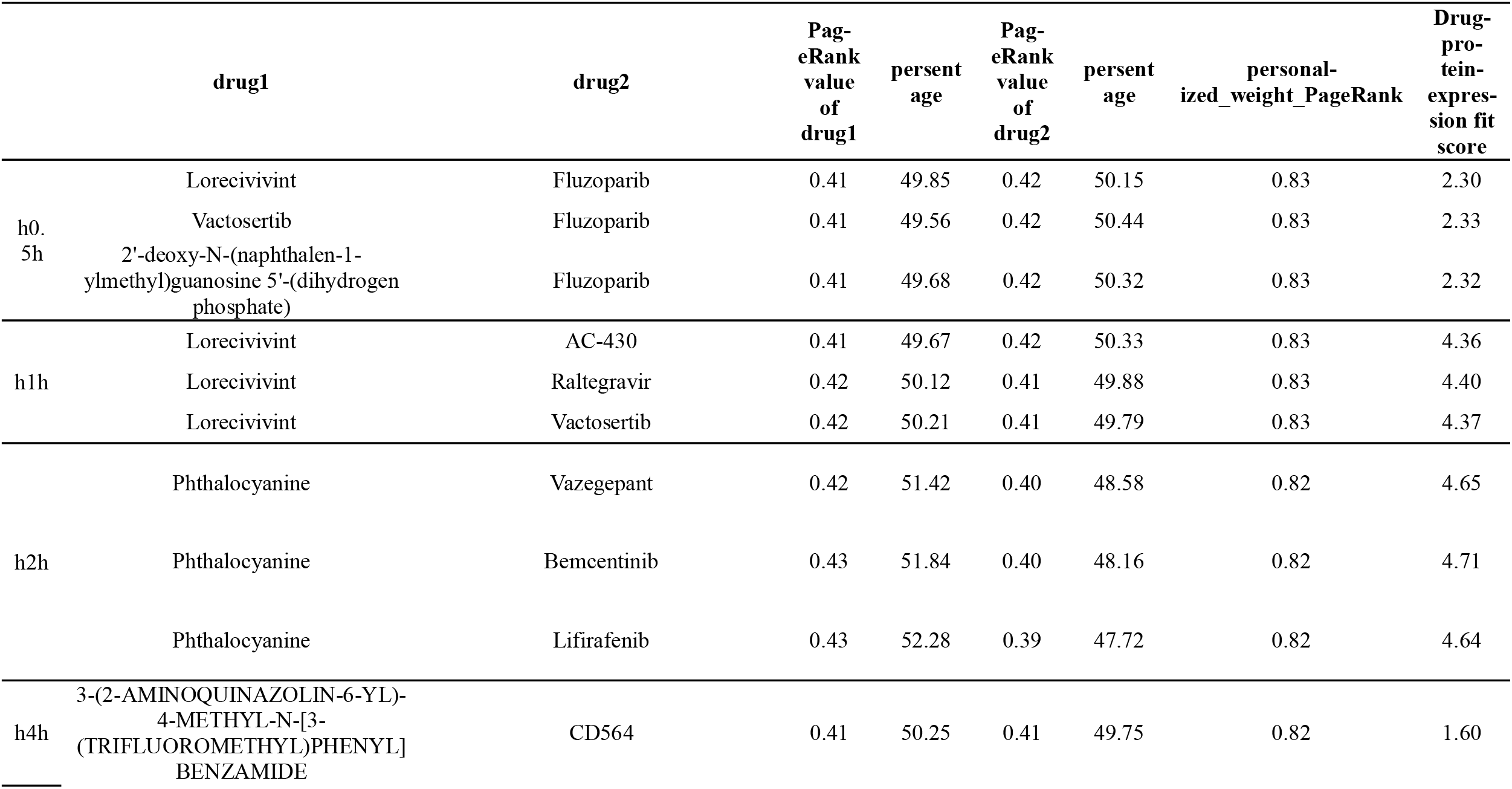
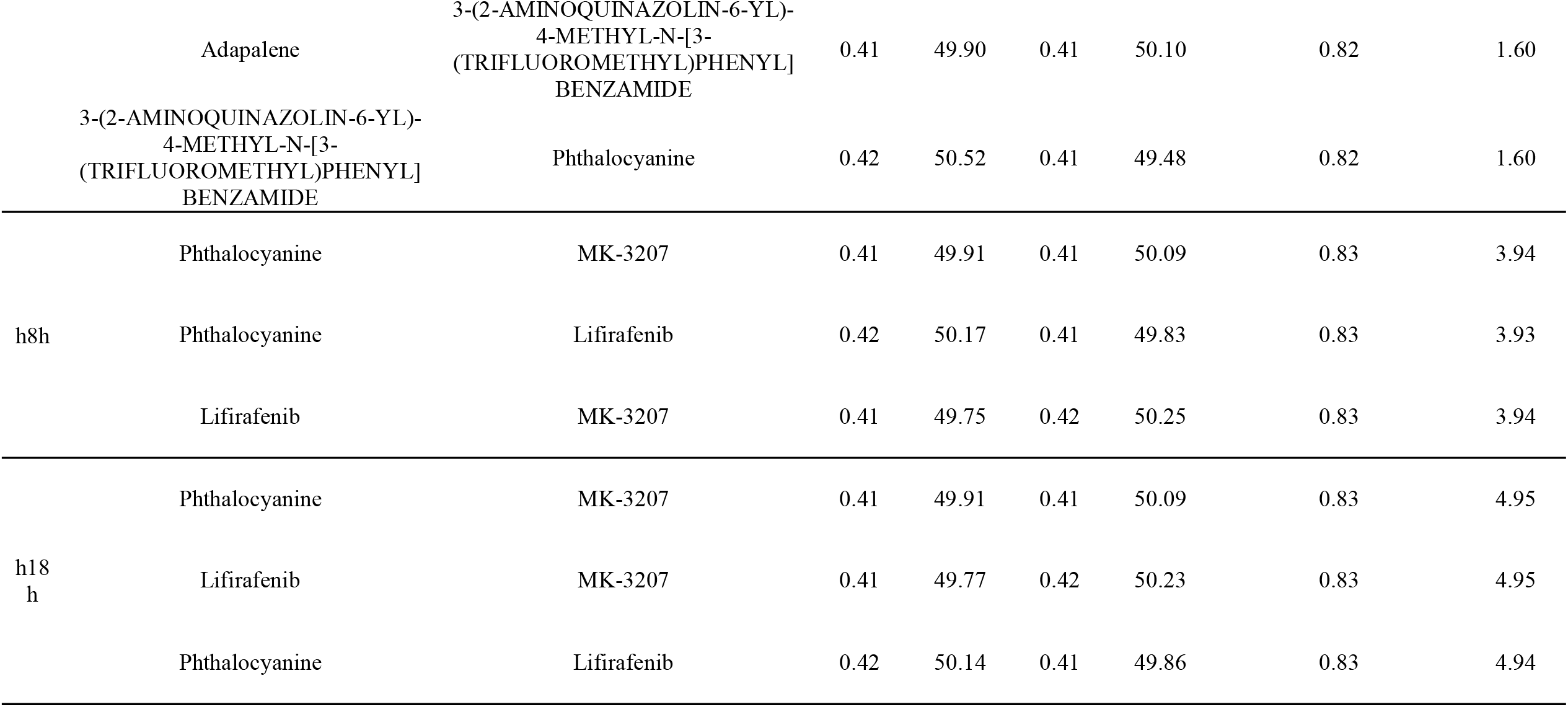
Rank of 2 drug combinations of different group.

## Discussion

In this report, a chronopharmacology-friendly multi-target drug discovery method is proposed by taking TH as example. This is the first application of PageRank and first attempt of AI protein prediction in the field of multi-target drug discovery.

In this paper, due to the high compatibility of PageRank, except for basic mRNA expression differences, the analysis of Weighted Gene Coexpression Network Analysis (WGCNA)[45] or Gene Regulatory Networks (GRN)[46] will be better because they could provide numerical results of all proteins. For the same reason, PageRank has good compatibility with existing pharmacological techniques such as pharmacophore models[47] and Quantitative Structure-Activity Relationship (QSAR)[48].

AlphaFold2 and RoseTTAFold were used for protein structure prediction. And the number of selected proteins predicted by AlphaFold2 in this research was close to that of RoseTTAFold. During the process of protein structure prediction, we found that for some proteins, the structures predicted by RoseTTAFold have less irregular structure than that of AlphaFold2. This may be due to the 2D distance map level being transformed and integrated by RoseTTAFold during neural network training[17], while AlphaFold2 only paired structure database and genetic database[16]. We also find a phenomenon that the predicted protein structures were relatively unstable under molecular dynamics simulation than preview reports of other protein structures detected by X-ray.

We use the PageRank algorithm to rank all the drugs and combinations. The application of PageRank is suitable. The combination of drug molecules is a memoryless stochastic process, which meets the qualifications of the Markov process. The comprehensive analysis involves free docking of proteins with all drugs. Drug combination analysis is to put proteins and related drugs into the solution for docking. Besides, the method has good compatibility with the wide compatibility of PageRank. In theory, all the technologies with numerical results can be ranked by the method. In addition, due to the charastic of PageRank, we can adjust the weight of different proteins to suppress the negative proteins with minimal impact on the positive ones. This property is very beneficial for chronopharmacology. It is difficult for one administration to affect the effect of the next one.

The new method brings new ideas for clinical drug development. For tumors, it is possible to effectively antagonize most of abnormally expressed proteins while limiting the affection of normal proteins. Given this approach’s close association with bioinformatics, bioinformatics can predict the most suitable drug for every patient in most diseases.

This research has some defects. 1: For the speed of calculating, we only choose the top 3 mRNAs for docking and the top 1 complex for MD simulation. Furthermore, the duration of molecular dynamics simulation is set to 50ns. These operations mitigate the rationality of the results relatively; 2. Theoretically, pharmacophore modeling has a better improvement under PageRank. But considering lacking related copyright and the purpose of the article, we chose to dock FBI-approved drugs with target proteins to explore the interaction between proteins and drugs.

In summary, this paper proposes a new method through protein structure predicting and PageRank. The results provide medical clues for the treatment of TH. This method takes a new attempt at drug discovery, which might make a little bit of a difference in pharmacology.

## Data Availability

https://github.com/FeiLiuEM/PageRank-weight-drug

## Acknowledge

We thank Zaizai Cao, Xiangjie Lin, and Yuanyuan Hao for the algorithm discussion.

